# Prospective cohort study of workers diagnosed with COVID-19 and subsequent unemployment

**DOI:** 10.1101/2021.07.05.21260053

**Authors:** Tomohisa Nagata, Masako Nagata, Ayako Hino, Seiichiro Tateishi, Akira Ogami, Mayumi Tsuji, Shinya Matsuda, Yoshihisa Fujino, Koji Mori, for the CORoNaWork project

## Abstract

**Objectives:** The purpose of this study was to investigate the relationships of workers being diagnosed with coronavirus disease 2019 (COVID-19) and being identified as close contacts of infected persons with unemployment in Japan.

**Methods:** This was a prospective cohort study using questionnaires about COVID-19 administered to Japanese workers. A baseline survey conducted on December 22–25, 2020, was used to determine history of being diagnosed with COVID-19 or being identified as a close contact of an infected person. Unemployment since the baseline survey was ascertained with a follow-up survey on February 18 and 19, 2021. The odds ratios of unemployment were estimated using a multilevel logistic model with adjusted covariates nested in prefecture of residence.

**Results:** Women (*n* = 8,771) accounted for 44% of the total sample (*n* = 19,941), and the mean age was 48.0 years. In terms of unemployment because of negative reasons, the multivariate analysis showed that the odds ratio of unemployment associated with being diagnosed with COVID-19 was 2.18 (95% CI: 1.05–4.55) and that the odds ratio associated with being identified as a close contact was 0.93 (95% CI: 0.29–2.95).

**Conclusions:** There is an association between workers being diagnosed with COVID-19 and unemployment. Occupational health professionals should follow up closely with workers diagnosed with COVID-19 after they return to work to prevent them from becoming unemployed against their will.

## Introduction

The spread of coronavirus disease 2019 (COVID-19) has become a global issue. The COVID-19 pandemic has had a profound impact not only on people’s daily lives, but also on the economy. The introduction of strong restrictive measures such as lockdowns in response to the pandemic has influenced the global economy, including by leading to an increase in unemployment rates worldwide.^1^ In Japan, the total unemployment rate gradually increased as COVID-19 spread, reaching 2.9% for men and women together in February 2021.^2^

Patients diagnosed with COVID-19 and individuals identified as close contacts of infected persons are isolated from those who are not sick. When these isolated people are workers, they are not able to go to work for a certain period of time. In Japan, workers with COVID-19 cannot go to work until at least 10 days have passed since symptom onset, at least 72 hours have passed since resolution of fever, and there has been an improvement in other symptoms.^3^ However, it is unclear how the subsequent continued employment of workers who are diagnosed with COVID-19 or identified as close contacts of infected persons is.

The purpose of this study was to investigate the relationship between workers being diagnosed with COVID-19 or identified as close contacts of infected persons and unemployment in Japan.

## Materials and methods

This prospective cohort study about COVID-19 among Japanese workers was conducted under the CORoNaWork (Collaborative Online Research on the Novel-coronavirus and Work) Project. The details of the study protocol have been described elsewhere.^4^ Briefly, we administered a baseline questionnaire on December 22–25, 2020, and a follow-up questionnaire on February 18 and 19, 2021, when Japan was in its third wave of the pandemic.

For the baseline survey, on December 22–25, 2020, a total of 33,087 workers were recruited throughout Japan from 605,381 randomly selected panelists who were registered with an Internet survey company. The inclusion criteria for participants were currently working and being aged 20–65 years. This study did not invite healthcare professionals or caregivers to participate. We used cluster sampling with stratification by sex, job type, and region. We excluded 6,051 invalid responses because of response time < 6 minutes, body weight < 30 kg or height < 140 cm, inconsistent answers to similar questions, and incorrect answers to questions intended to identify fraudulent responses. We distributed the follow-up questionnaire to the 27,036 people with valid responses to the baseline questionnaire on February 18 and 19, 2021. In total, 19,941 participants completed both questionnaires (follow-up rate = 73.8%).

The present study was approved by the Ethics Committee of the University of Occupational and Environmental Health, Japan (reference No. R2-079 and R3-006). Informed consent was obtained from all participants.

### Explanatory variables

The baseline survey asked two questions about the participants’ COVID-19 diagnostic history. First, the participants were asked, “Have you had COVID-19?” with the response options of *yes* or *no*. If the answer to this question was *yes*, the respondent was considered a worker being diagnosed with COVID-19. If the answer to the question was *no*, to determine whether they had been identified as a close contact of an infected person, the respondent was asked, “Have you been identified as a close contact of a person diagnosed with COVID-19?” with the response options of *yes* or *no*. Workers answering *yes* to this question were identified as close contacts. Workers reporting that they had been diagnosed with COVID-19 or identified as close contacts should have been classified in this way according to public health center instructions, so we believe that these questions allowed us to identify infected and exposed people with a high degree of accuracy.

### Outcomes

Unemployment was ascertained in the following way. First, the baseline survey included only people who were employed at the time of response. In the follow-up survey, in response to the question “Have you changed your place of work since December 2020?” respondents were asked to select one of the following six options: *no change, I was transferred to another company, I resigned and got a new job right away, I stopped working and was unemployed for a period of time but am now working, I stopped working and started a business (e*.*g*., *managing a company, running a sole proprietorship, or engaging in self-employment)*, and *I stopped working and am currently not working (including job seeking)*. We wanted to identify unemployment because of negative reasons as the primary outcome to achieve the research objectives, so we defined unemployment using the responses of *I stopped working and was unemployed for a period of time but am now working* and *I stopped working and am currently not working (including job seeking)*. As a secondary outcome, we also considered unemployment regardless of the reason, comparing respondents who chose *no change* or *I was transferred to another company* with those who chose any other option.

### Control variables

We retrieved the following data from the baseline survey for inclusion as control variables: age, sex, annual household income, educational attainment, marital status, job type, self-rated health, and region.

Age was treated as a continuous variable. Education was classified into five as follow: junior high school, high school, junior college or technical school, university, and graduate school. Yearly household income was classified into four as follow: less than 2.50 million Japanese yen (JPY), 2.50–3.75 million JPY, 3.75–5.25 million JPY, and greater than 5.25 million JPY. Marital status was classified into three as follow: married, divorced or widowed, and never married. Job type was classified into three categories: mainly desk work, work mainly involving interpersonal communication, and mainly physical work. The following question was asked to assess self-rated health: “Please choose the option that best describes your overall health condition, including both physical and mental health” with the following five response options: *extremely good, very good, good, fair*, and *bad*.

In addition, prefecture of residence was included as a community-level variable.

### Statistical analysis

The odds ratios (ORs) of unemployment (primary and secondary outcome) for infected workers and persons identified as close contacts of infected persons were estimated using a multilevel logistic model nested in prefectures of residence. The multivariate model was adjusted for age and sex, educational attainment, annual household income, marital status, job type, and self-rated health. The incidence rate of COVID-19 by prefecture was included as a prefecture-level variable. *P*-values less than 0.05 were considered statistically significant. All analyses were conducted using Stata (Stata Statistical Software release SE16.1; StataCorp LLC, College Station, TX, USA).

## Results

The basic characteristics of the respondents are shown in Table 1. There were 8,771 women in the sample, accounting for 44% of the total (*n* = 19,941). The mean age was 48.0 years, with 454 (2.3%) reporting experiencing unemployment because of negative reasons and 725 (3.6%) reporting experiencing unemployment for any reason.

**Table 1.** Basic characteristics of study respondents

|  | Workers who<br>were not infected | Workers<br>identified as<br>close contacts<sup>a</sup> | Workers<br>diagnosed with<br>COVID-19 |
| --- | --- | --- | --- |
| Number of respondents | 19,646 | 141 | 154 |
| Age, mean (SD) | 48.0 (10.1) | 45.7 (10.6) | 43.8 (11.5) |
| Men | 10,998 (56.0%) | 87 (61.7%) | 85 (55.2%) |
| Education |  |  |  |
| Junior high or high school | 5300 (27.0%) | 23 (16.3%) | 37 (24.0%) |
| Vocational school, junior college, or technical school | 4513 (23.0%) | 37 (26.2%) | 35 (22.7%) |
| University | 8742 (44.5%) | 70 (49.6%) | 68 (44.2%) |
| Graduate school | 1091 (5.6%) | 11 (7.8%) | 14 (9.1%) |
| Annual income (million JPY) |  |  |  |
| < 2.50 | 4183 (21.3%) | 17 (12.1%) | 33 (21.4%) |
| ≥ 2.50 and < 3.75 | 5388 (27.4%) | 34 (24.1%) | 36 (23.4%) |
| ≥ 3.75 and < 4.99 | 4750 (24.2%) | 35 (24.8%) | 36 (23.4%) |
| ≥ 4.99 | 5325 (27.1%) | 55 (39.0%) | 49 (31.8%) |
| Marital status |  |  |  |
| Married | 11,004 (56.0%) | 85 (60.3%) | 96 (62.3%) |
| Widowed/divorced | 2012 (10.2%) | 10 (7.1%) | 14 (9.1%) |
| Unmarried | 6630 (33.7%) | 46 (32.6%) | 44 (28.6%) |
| Job type |  |  |  |
| Mainly desk work | 10,135 (51.6%) | 61 (43.3%) | 72 (46.8%) |
| Work mainly involving interpersonal communication | 4847 (24.7%) | 44 (31.2%) | 46 (29.9%) |
| Mainly physical work | 4664 (23.7%) | 36 (25.5%) | 36 (23.4%) |
| Self-rated health |  |  |  |
| Extremely good | 542 (2.8%) | 6 (4.3%) | 22 (14.3%) |
| Very good | 1938 (9.9%) | 19 (13.5%) | 35 (22.7%) |
| Good | 7374 (37.5%) | 50 (35.5%) | 42 (27.3%) |
| Fair | 7270 (37.0%) | 49 (34.8%) | 45 (29.2%) |
| Bad | 2522 (12.8%) | 17 (12.1%) | 10 (6.5%) |
| Unemployment |  |  |  |
| Unemployment because of negative reasons | 443 (2.3%) | 3 (2.1%) | 8 (5.2%) |
| Unemployment regardless of the reason | 700 (3.6%) | 6 (4.3%) | 19 (12.3%) |

<sup>a</sup> Workers identified as close contacts of people who were diagnosed with COVID-19
Abbreviations: JPY, Japanese Yen

Table 2 shows the associations of being diagnosed with COVID-19 and being identified as a close contact of someone with COVID-19 with unemployment. For the primary outcome of unemployment because of negative reasons, the multivariate analysis showed that the OR of unemployment associated with being diagnosed with COVID-19 was 2.18 (95% CI: 1.05–4.55) and that the OR associated with being identified as a close contact was 0.93 (95% CI: 0.29–2.95).

**Table 2.**
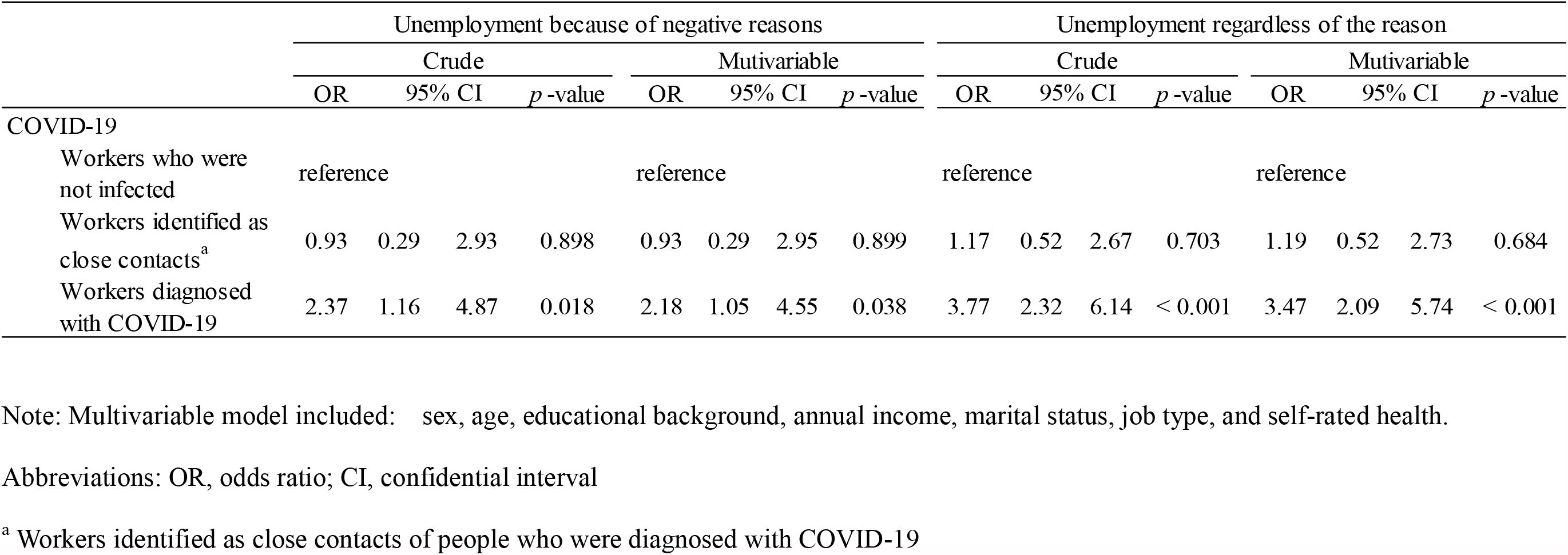
Associations of being diagnosed with COVID-19 and being identified as a close contact of an infected person with unemployment

For the secondary outcome (unemployment regardless of the reason), the multivariate analysis showed that the OR of unemployment associated with being diagnosed with COVID-19 was 3.47 (95% CI: 2.09–5.74) and that the OR associated with being identified as a close contact was 1.19 (95% CI: 0.52–2.73).

## Discussion

We evaluated the relationships of workers having COVID-19 and being identified as close contacts with unemployment. We observed an association between workers being diagnosed with COVID-19 and unemployment but not between being identified as close contacts and unemployment.

There are several possible reasons why workers diagnosed with COVID-19 subsequently became unemployed. First, workers with COVID-19 may have recovered once but then been unable to work as a result of suffering from the long-term effects of COVID-19. A recent longitudinal prospective cohort study of post-COVID-19 patients found that 30% had at least one symptom persisting for nine months or longer.^5^ The sequelae of COVID-19 are referred to as “post-COVID syndrome” or “COVID long haul syndrome.”^6^ However, in our analysis, the relationship between being diagnosed with COVID-19 and unemployment remained significant even after adjusting for self-rated health; thus, poor health may not be the reason for unemployment.

Second, workers who had COVID-19 may have resigned as a result of feeling uncomfortable in the workplace. Workers with COVID-19 have to be away from work for about two weeks. If coworkers are in close contact with the infected people, the coworkers must also take time off work. Workers with COVID-19 may have felt bad about having a major impact on their workplace and therefore have resigned. This situation may also have led to bullying and harassment in the workplace. Such harassment has been reported to be a common occurrence among healthcare workers^7^, but it can also occur in the general workforce. Workplace bullying, in addition to increasing mental distress and even work-related suicide, also has socioeconomic consequences related to sick leave and unemployment.^8^ Although workers who were identified as close contacts of infected people also needed to take a certain amount of time off work, they did not spread the infection to others in the workplace; this may have prevented workers identified as close contacts from being forced into resigning.

Some limitations of this study warrant mention. First, exposure was assessed by asking about the existence of a history of COVID-19 in the baseline survey (at the end of December 2020), but it was not clear when respondents had been diagnosed with COVID-19 from January 16, 2020 (when the first case of COVID-19 was reported in Japan) and December 2020 (when the baseline survey was conducted). However, the number of people in Japan diagnosed with COVID-19 was overwhelmingly higher after October 2020, so it is very likely that most respondents in this study who had COVID-19 were diagnosed from October to December 2020. Second, information on past diagnosis was self-reported. However, almost all COVID-19 patients and people identified as close contacts are diagnosed after undergoing polymerase chain reaction testing. This process is rigorously followed to prevent the spread of infection, so the information is likely to be fairly accurate. Third, we did not investigate the reasons for unemployment except for starting their own business. As a result, we were unable to fully distinguish between cases in which workers resigned voluntarily and those in which their employment was terminated against their will. Future research should conduct analyses using survey data that allow for a thorough assessment of the reasons for unemployment.

In conclusion, there is an association between workers being diagnosed with COVID-19 and unemployment. Occupational health professionals need to follow up closely with workers diagnosed with COVID-19 after their return to work to prevent them from becoming unemployed against their will.

## Data Availability

The data that support the findings of this study are available from the corresponding author, Tomohisa Nagata, upon reasonable request.

## Acknowledgments

This study was supported and partly funded by the University of Occupational and Environmental Health, Japan; General Incorporated Foundation (Anshin Zaidan); The Development of Educational Materials on Mental Health Measures for Managers at Small-sized Enterprises; Health, Labour and Welfare Sciences Research Grants; Comprehensive Research for Women’s Healthcare (H30-josei-ippan-002); Research for the Establishment of an Occupational Health System in Times of Disaster (H30-roudou-ippan-007), scholarship donations from Chugai Pharmaceutical Co., Ltd., the Collabo-Health Study Group, and Hitachi Systems, Ltd.

The current members of the CORoNaWork Project, in alphabetical order, are as follows: Dr. Yoshihisa Fujino (present chairperson of the study group), Dr. Akira Ogami, Dr. Arisa Harada, Dr. Ayako Hino, Dr. Hajime Ando, Dr. Hisashi Eguchi, Dr. Kazunori Ikegami, Dr. Kei Tokutsu, Dr. Keiji Muramatsu, Dr. Koji Mori, Dr. Kosuke Mafune, Dr. Kyoko Kitagawa, Dr. Masako Nagata, Dr. Mayumi Tsuji, Ms. Ning Liu, Dr. Rie Tanaka, Dr. Ryutaro Matsugaki, Dr. Seiichiro Tateishi, Dr. Shinya Matsuda, Dr. Tomohiro Ishimaru, and Dr. Tomohisa Nagata. All members are affiliated with the University of Occupational and Environmental Health, Japan.

## Disclosure

### Ethical approval

This study was approved by the ethics committee of the University of Occupational and Environmental Health, Japan(reference No. R2-079 and R3-006).

### Informed Consent

Informed consent was obtained in the form of the website.

### Registry and the Registration No. of the study/Trial

N/A

### Animal Studies

N/A

